# Systematically omitting indoor air quality: sub-standard guidance for shelters, group homes and long-term care during the COVID-19 pandemic

**DOI:** 10.1101/2022.01.26.22269908

**Authors:** Amy S. Katz, Tianyuan Li, LLana James, Jeff Siegel, Patricia O’Campo

## Abstract

Scientific evidence suggests that COVID-19 is transmitted through the air, and can be mitigated using indoor air quality measures such as ventilation and filtration. We set out to explore how Public Health Ontario (PHO) responded to this evidence. PHO is mandated by legislation to share “scientific and technical advice and support” and “contribute to efforts to reduce health inequities” in Ontario, Canada, a jurisdiction of more than 14 million people (Ontario Agency for Health Protection and Promotion Act, 2007). We were particularly interested in how PHO addressed indoor air quality measures in the context of guidance developed for communal facilities such as long-term care homes and congregate settings (e.g. shelters, group homes, detention centres). We reviewed PHO’s public, written COVID-19 guidance specifically designed for long-term care and congregate settings, and published or revised between January, 2021 and October, 2021. We also compared PHO’s COVID-19 checklists for long-term care and congregate settings to PHO’s COVID-19 checklists for schools, camps and doctor’s offices. We found no references to indoor air quality measures in any guidance documents specifically designed for long-term care and congregate settings, including COVID-19 checklists. We did, however, find references to indoor air quality measures in COVID-19 checklists for schools, summer camps and doctor’s offices. We conclude that PHO has provided sub-standard COVID-19 guidance to long-term care and congregate settings, putting workers and residents at greater risk of illness and death, and exacerbating health inequities in Ontario.

## Introduction

In 2006, the SARS Commission completed its investigation into the 2003 outbreak of SARS in the province of Ontario, Canada, which killed 44 people (see references to jurisdiction). At its conclusion, the Commission made one central recommendation: Ontario’s public health and health care systems should adopt the precautionary principle in the face of future pandemics, particularly in the context of airborne transmission. In his final summary report, Commission lead Archie Campbell wrote:

> Those who argued against the N95, which protects against airborne transmission, believed SARS was spread mostly by large droplets. As a result, they said, an N95 was unnecessary except in certain circumstances and a surgical mask was sufficient in most instances. They made this argument even though knowledge about SARS and about airborne transmission was still evolving. That more and more studies have since been published indicating the possibility under certain circumstances of airborne transmission, not just of SARS but of influenza, suggests the wisdom and prudence of taking a precautionary approach in the absence of scientific certainty.^1,p11^

Campbell referenced studies that indicate airborne transmission of both SARS and influenza. These included a study that determined that airborne transmission was responsible for a SARS outbreak in a housing complex, and one that detected the SARS virus in the air in a hospital room.^2,3^ In terms of influenza, Campbell cited a review published by Centers for Disease Control that concludes that, “Published evidence indicates that aerosol transmission of influenza can be an important mode of transmission…”^4^

In 2008, in part to address the failures of Ontario’s public health system during the SARS outbreak, Ontario established a new agency, now called Public Health Ontario (PHO).^5^ PHO is mandated by legislation to “enhance the protection and promotion of the health of Ontarians and to contribute to efforts to reduce health inequities” through the provision of “scientific and technical advice and support.” The organization has particular responsibilities in the context of respiratory pandemics, including, “evaluating the modes of transmission of febrile respiratory illnesses…”^5,6^

Twelve years later, in March, 2020, the World Health Organization declared the COVID-19 outbreak a pandemic (see Figure 1). Within months, scientists, engineers and physicians began urging public health authorities to address airborne transmission. In July, 2020, 239 scientists (including a co-author on this paper) endorsed a commentary in the journal *Clinical Infectious Diseases* appealing “to the medical community and to the relevant national and international bodies to recognize the potential for airborne spread of coronavirus disease 2019…”^7^ In November, 2020, Ontario-based physicians and engineers wrote to the Ontario Minister of Health and Long-term Care, the Ontario Chief Medical Officer of Health, and the Vice-President of PHO, urging them to, “update the province’s COVID-19 guidelines, regulations and public communication to reflect the importance of ventilation…”^8^ This was followed in January, 2021, with an open letter from scientists, physicians and engineers across Canada demanding public health measures such as ventilation assessments, portable HEPA filters, CO2 monitoring, and fit-tested N95 masks for health care and other essential workers.^9^

**Figure 1.**
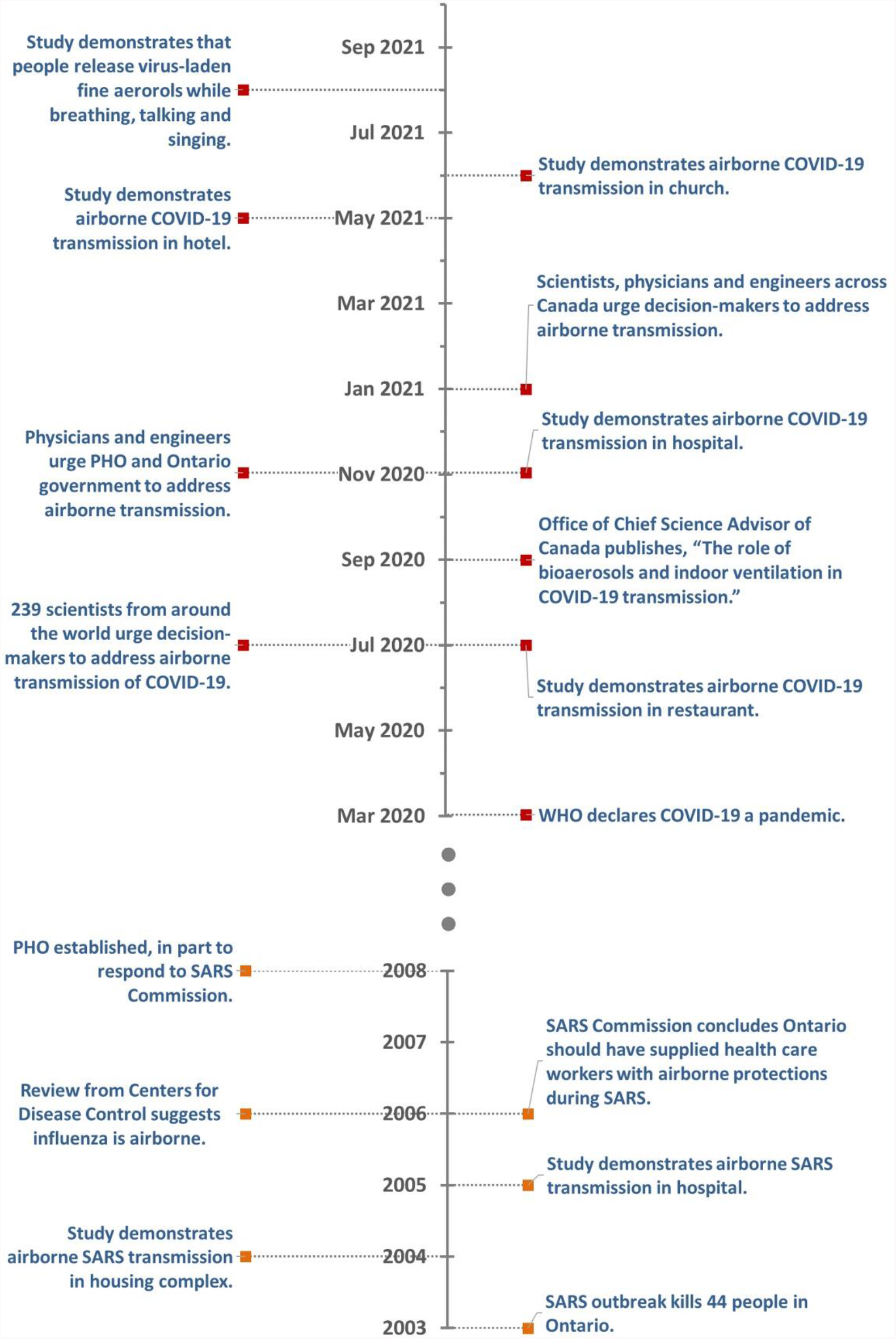
SARS and COVID-19 timeline in Ontario.

Expert reports and warnings were accompanied by peer-reviewed evidence demonstrating that COVID-19 spreads through the air. These include studies documenting airborne transmission in settings such as hospitals, restaurants, churches and hotels, and those demonstrating that people infected with COVID-19 will release fine aerosols when breathing, talking or singing.^10–15^ In addition, some scientists explored the dynamics of the COVID-19 pandemic itself, or of respiratory viruses in general, to demonstrate that COVID-19 is airborne.^16,17^

At the same time, scientists and engineers published guidance on the use of indoor air quality (IAQ) measures to mitigate airborne transmission of COVID-19. ^18–21^ These measures include increasing ventilation and filtration through measures such as optimizing HVAC systems; portable HEPA filters; bathroom fans that exhaust to the outside; and natural ventilation such as windows. Additional IAQ measures include upper-room and in-duct ultra-violet disinfection, and protocols to clear the air in rooms between groups of people.

In October, 2021, we set out to explore how PHO responded to: evidence related to airborne transmission of COVID-19, SARS and influenza; the potential for IAQ measures to mitigate harm; and the SARS Commission’s recommendation to adopt the precautionary principle. We were particularly interested in how PHO addressed IAQ measures in the context of guidance developed for institutional settings where large numbers of people live communally, often termed “congregate settings” in Ontario.

The health and safety measures recommended to congregate settings have particular implications for health equity in Ontario. The state and its institutions, by way of inter-related structures such as colonization, white supremacy and ableism, determine the populations that are compelled to live in shelters, detention centres, group homes and long-term care, and the populations that are generally able to avoid these same facilities. As a result, and given PHO’s legislative mandate to “contribute to efforts to reduce health inequities,” particular scrutiny should be given to the quality of the guidance shared with these facilities (see “health equity and human rights frameworks in Ontario”).

In addition, it is clear that institutions with communal living conditions face particularly high risks. To date, more than 4,000 long-term care home residents and 10 workers have died of COVID-19, constituting approximately 40 per cent of COVID-19 deaths in Ontario.^22^ Some long-term care homes have lost more than 30 per cent of their residents to the pandemic.^23^ Beyond long-term care homes, we know of no sources of systematic, province-wide data broken down by type of facility and including both case counts and deaths. However, reports from mainstream media, independent journalists and academic researchers suggest that outbreaks in facilities such as shelters, correctional facilities and group homes for adults with disabilities have been severe.^24–26^

PHO is mandated by legislation to share “scientific and technical advice and support” and “contribute to efforts to reduce health inequities” in Ontario, a jurisdiction of more than 14 million people.^6^ We chose to explore PHO’s guidance for long-term care and congregate settings during the COVID-19 pandemic. To do this, we conducted a review of all written guidance specifically designed for long-term care and congregate settings and posted on PHO’s website in late October, 2021. As we were interested in how PHO responded to warnings and evidence related to airborne transmission and indoor air quality, we focused on resources published on or after January 1, 2021, a point at which the potential for COVID-19 to spread through the air was well-known (see Figure 1).

## Definitions and context

### References to jurisdiction

While Canadian jurisdictions are often referred to as fixed and legitimate categories, they are the product of land theft from Indigenous nations and groups. Ownership and/or use by Canada is often asserted in contradiction to Canada’s own laws and treaty obligations, and the original agreements between Indigenous nations and groups and the British Crown. In addition, there are wide swathes of territory for which there are no treaties or agreements, and to which Canada has no claim at all. Finally, Canadian jurisdictions are not contiguous—the land labelled as Canada on most maps is not all under Canadian jurisdiction. It would be inaccurate to refer to settler jurisdictional names and boundaries in Canada without pointing to the fact that they are imposed, relatively recent and, in many cases, contested and in flux. We do not refer to jurisdictions such as Ontario to suggest these are fixed and inevitable categories. Rather, we explore the mandate and actions of PHO in the context of the current settler-colonial matrix of governance that helps to determine the conditions under which many people in what is currently called Ontario work and live.

### History of PHO

In 2003, Ontario had one of the largest outbreaks of SARS in the world. Forty-four people died what many saw as preventable deaths.^1^ In 2006, the SARS Commission released its final report, “based on public hearings, government and hospital documents, and confidential interviews of more than 600 people connected with SARS…”^1p.3^ The report underlined the need to address Ontario’s fragmented, under-resourced public health system. It also found that the public health and health care systems had not applied the “precautionary principle” during the SARS outbreak—they had failed to err on the side of caution when it came to health and safety, in particular in the context of potential airborne transmission.

In part to respond to the public health failures identified by the SARS Commission, Ontario established a new agency “to provide scientific and technical advice in the areas of health protection and promotion.”^5p.267^ Public Health Ontario (PHO), originally called the Ontario Agency for Health Protection and Promotion, was established in 2008 following the passing of the *Ontario Agency for Health Protection and Promotion Act*. The Act tasks PHO with particular responsibilities in the context of infectious disease, and respiratory outbreaks in particular. These include: informing provincial policy related to infection prevention and control; providing “scientific and technical advice and operation supports” during outbreaks; and “evaluating the modes of transmission of febrile respiratory illnesses…”^1^

### Health equity and human rights frameworks in Ontario

On its web page, “health equity and COVID-19,” PHO shares the following introductory text:

> Racial and socioeconomic inequities in rates of COVID-19 infection, hospitalization and mortality have been identified in Canada and internationally. Social determinants of health (SDOH), such as gender, socioeconomic position, race/ethnicity, occupation, Indigeneity, homelessness and incarceration, are factors that potentially increase risk and severity of COVID-19 infection. Incorporating SDOH into risk considerations and assessments is crucial for supporting an equitable COVID-19 response.^27^

Many have pointed out, however, that the factors that drive preventable ill health are not to be found in individuals or populations, but in structures such as colonization and white supremacy that systematically produce negative outcomes for some groups, ^28,29,30,31^ and favourable outcomes for others.^32^ Researchers have pointed out that framings such as “health equity,” “social determinants of health,” “vulnerable,” and “underserved” are broad and vague, and therefore can serve to obscure the role of power structures and their attendant institutional violence in producing preventable ill health.^31,33^

Despite its conceptual malleability, “health equity” can be useful when applied strategically, and with precision, in particular as it is mandated in different ways across social services, health care and public health in Ontario. For example, the provincial legislation that created PHO tasks the organization with contributing, “to efforts to reduce health inequities.”^6^ The Ontario Public Health Standards, which apply to all local Boards of Health, enshrine the following goal:

> Public health practice results in decreased health inequities such that everyone has equal opportunities for optimal health and can attain their full health potential without disadvantage due to social position or other socially determined circumstances.^34,p23^

From a legal standpoint, the Ontario Human Rights Code prohibits discrimination based on “protected grounds” which include factors such as age; ancestry, colour and race; citizenship and disability. The Code applies to specific areas of society, and enshrines, “equal treatment with respect to services, goods and facilities,” including services provided by provincial and municipal governments such as long-term care homes and congregate settings. ^35,36^

### How PHO defines congregate settings

On its COVID-19 web page titled “COVID-19 resources for congregate living settings,” PHO defines congregate facilities as follows:

Congregate living settings refer to a range of facilities where people (most or all of whom are not related) live or stay overnight and use shared spaces (e.g., common sleeping areas, bathrooms, kitchens) including:

- Shelters
- Group homes
- Correctional facilities
- Children or youth residential settings

Public Health Ontario has developed a variety of resources to help staff and administrators address COVID-19 prevention and control in these settings. These resources can be used along with the support provided by local public health units. For resources related to COVID-19 in long-term care and retirement homes, please visit our Long-Term Care Resources page.^37^

As above, in most of its COVID-19 guidance, PHO draws a distinction between general congregate settings and long-term care homes. In some cases, long-term care homes are also listed as health care settings. In others, they are bundled with retirement homes. Finally, while much of the guidance for congregate settings is framed by PHO as applying to congregate settings in general (excluding long-term care homes), some guidance documents state that they are not specifically developed for correctional facilities.

## Methods

### Thematic analysis and document review

We identified, stored and analyzed documents using largely standard document review methods.^38^ Our research process was informed by thematic analysis (TA), which, like many qualitative methods, seeks to identify patterns within data sets. TA is marked, in part, by its emphasis on transparency, encouraging researchers to articulate assumptions and ensure that readers understand how the research was done.^39^ As a result, we have included a detailed methods section, along with tables detailing our research sample. We have also listed some of our core assumptions in the section below.

TA also directs researchers to contextualize data, rather than simply present findings. To this end, we have included detailed “introduction” and “definitions and context” sections in order to ensure that our findings are understood: a) in the context of our collective understanding of concepts such as “inequity;” and b) in local context (for example, legislative context). In addition, TA emphasizes the agency of the researcher, and discourages passive framings such as “themes emerged.”^39,p.80^ Rather, themes are a product of research decisions that should be articulated at every step. In response, we have attempted to explicate both our research decisions and conceptual understandings throughout this paper.

Finally, TA is an iterative process, moving back and forth across stages of data collection and analysis. Co-authors spent months visiting and revisiting every stage of the research as described below, and adding layers of context as a group.

We did not apply all stages of TA, and do not claim that this is a “TA manuscript.” Rather, our approach was informed by the method’s emphasis on iterative processes, transparency, researcher agency, and contextualization of research findings.

### Assumptions

As above, TA encourages researchers to articulate the assumptions that led to their research question, shaped their study design and informed “what can be claimed on the basis of [the] data” they are presenting.^40,p.337^ We share the assumption that public health measures should be informed by the precautionary principle, and err on the side of caution when it comes to health and safety. This is particularly true in the case of measures such as ventilation and filtration, where expert implementation generates few, if any, risks. We also share the assumption that there is a particularly strong imperative for public health to move aggressively to protect people who are compelled by the state to live in specific types of facilities. Finally, we share the assumption that, to some degree, written guidance from PHO has helped to influence the actions of those responsible for health and safety in long-term care and congregate settings throughout the COVID-19 pandemic.

### Document review process

#### Identifying and selecting documents

We explored written guidance listed in the COVID-19 “congregate living” and “long-term care” sections of PHO’s website as of October 29, 2021. We focused on documents published on or after January, 2021, as, by this time, the potential for IAQ measures to mitigate COVID-19 had been widely-discussed (see Figure 1). We then divided resources into three categories:

a) <u>General COVID-19 guidance developed specifically for long-term care and congregate settings</u>. We excluded documents that were not guidance documents, for example, epidemiological reports. We also excluded documents that were lists of other documents.

b) <u>Topic-specific COVID-19 guidance developed specifically for long-term care and congregate settings</u> (e.g. resources on personal protective equipment). Where a topic encompassed a range of potential mitigation measures (for example, cohorting) it was included in the category of “general guidance.”

c) <u>COVID-19 checklists</u>. In this section, we set out to compare guidance for long-term care and congregate settings with parallel guidance for other settings. To do this, we chose PHO’s COVID-19 “checklists,” which share a relatively standard format. Each checklist identifies its audience and purpose, and then provides an itemized checklist divided into categories. Each item on the checklist has a section for notes, or in which to mark “yes” or “no” beside a particular measure, presumably so that the person auditing the facility can check off measures that are in place, and identify measures that still need to be implemented. Some checklists also include signature areas for positions such as public health staff, inspectors, facility administrators or facility staff. To identify all relevant checklists, we used the search filter function on PHO’s website, filtering for “type of resource” (checklist) and “topic” (COVID-19).

See Figure 2 for a flow chart illustrating our approach to document selection.

**Figure 2.**
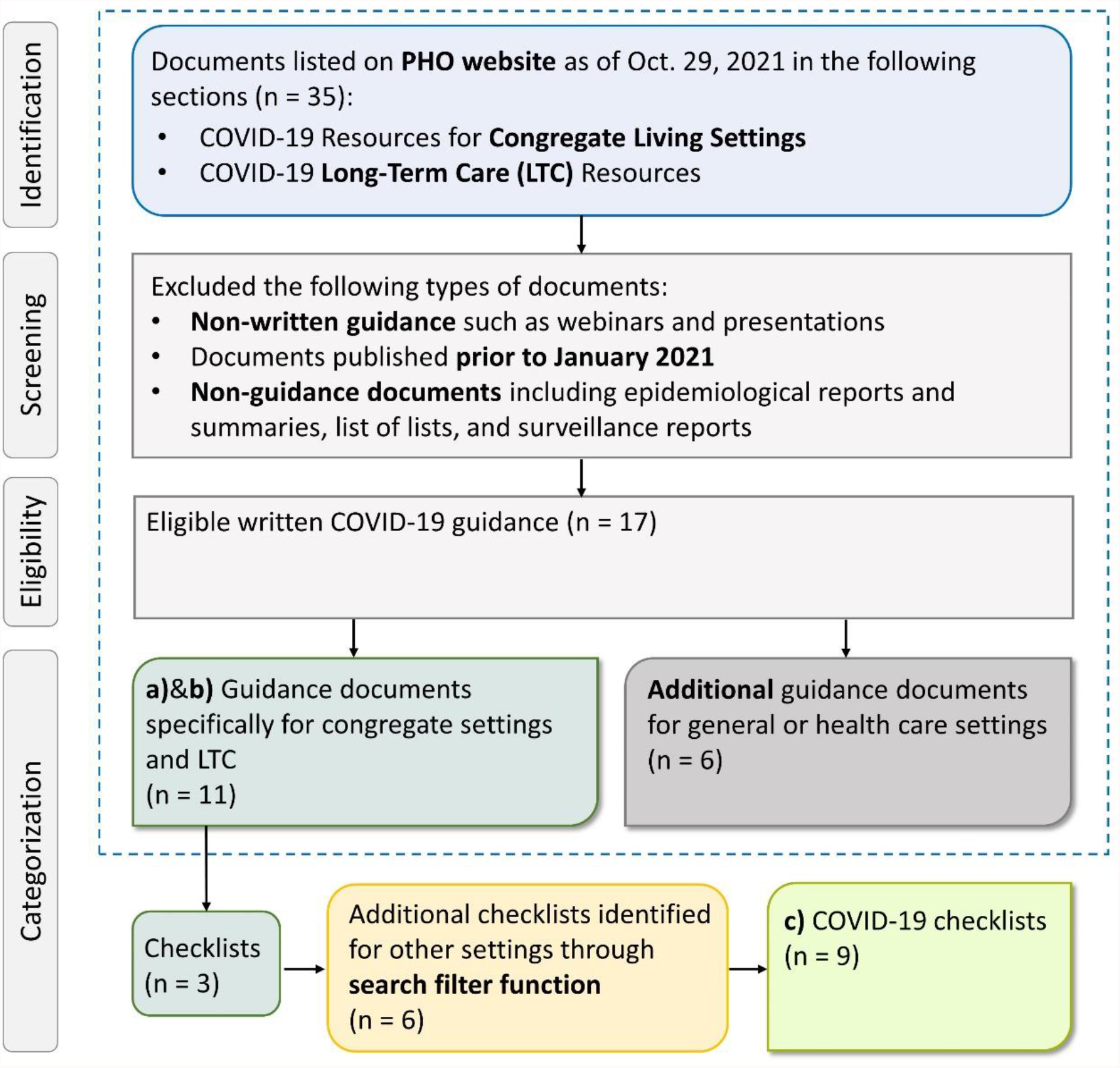
Flow chart illustrating document selection approach.

### Reviewing resources

#### Key word search

We searched each resource in the “general guidance” and “checklist” categories for the following key words, in order to explore PHO’s approach to indoor air quality: air, airborne, aerosol(s), HVAC, ventilation, ventilate, filtration, filter, HEPA, portable, exhaust, fan, window(s), ultraviolet and UV. We chose these keywords because best practices for reducing airborne transmission (also termed “aerosol” transmission) of COVID-19 through improvements to indoor air quality include ventilation, filtration, clearing the air in rooms between groups, and, in some cases, ultraviolet disinfection.^18–21^ We began to identify these keywords through our pilot rapid review of COVID-19 guidance recommended by Toronto Public Health.^41^

To conduct the search, we used the search function in Adobe PDF software, and selected the “stemming” option, which highlights all words that include any segment of the search term. For example, a search for “ventilation” will include “ventilate” and “ventilated.” We also read each document in this category to ensure we didn’t miss key words through our electronic search.

#### Qualitative review

We read each document in the “general guidance” and “checklists” category to explore its approach to indoor air quality in the context of COVID-19 mitigation. We were also interested in the question of which mitigation measures were emphasized. Finally, we conducted a careful reading of the “revisions” section of each document to identify changes to guidance over time.

We did not conduct detailed qualitative reviews for topic-specific guidance, since the titles generally made their focus clear (e.g. guidance on personal protective equipment).

### Storing resources

We stored archived web pages and PDF copies of documents both using web-based software and on an external hard drive.

## Findings

### Guidance designed for long-term care homes and congregate settings

We identified 11 COVID-19 guidance documents developed specifically for long-term care homes (n. 5) and congregate settings (n. 6). Of these, nine are general guidance documents and two are topic-specific guidance documents. Documents are in different formats, and content often overlaps. Document formats, as named by PHO, are as follows: “at a glance” (n. 4), “checklists” (n. 3), “best practice” (n.1), “environmental scan,” (n. 1), “infographic” (n.1) and “reference guide” (n. 1).

In the eleven documents in our sample, there are no references to IAQ measures. Instead, resources emphasize infection control and prevention measures such as: personal protective equipment, entrance screening, cohorting, surveillance and communication, vaccination, symptom-monitoring, hand hygiene, respiratory etiquette (such as covering mouth and nose with sleeve while sneezing), cleaning and disinfection, and physical distancing, including the use of physical barriers, floor markers or signage to keep people two metres apart.

Several resources emphasize the need for private bedrooms and bathrooms for people with confirmed or suspected cases of COVID-19. To mitigate transmission when private spaces are not available, some of these same resources suggest barriers such as curtains in bedrooms, sleeping head-to-toe, and cleaning and disinfecting bathrooms between cohorts.

Many resources stipulate the need for “droplet and contact” as opposed to “airborne” precautions. In Ontario, droplet and contact precautions include gloves, gowns, eye protection and surgical masks.^42^ They do not, however, include respirator-grade masks, or specific ventilation requirements such as negative pressure rooms.^a^

Of the eleven documents in our sample, six had undergone one or more revisions, some as recently as September 2021.

See Table 1 for a full list of resources included in this section.

**Table 1.**
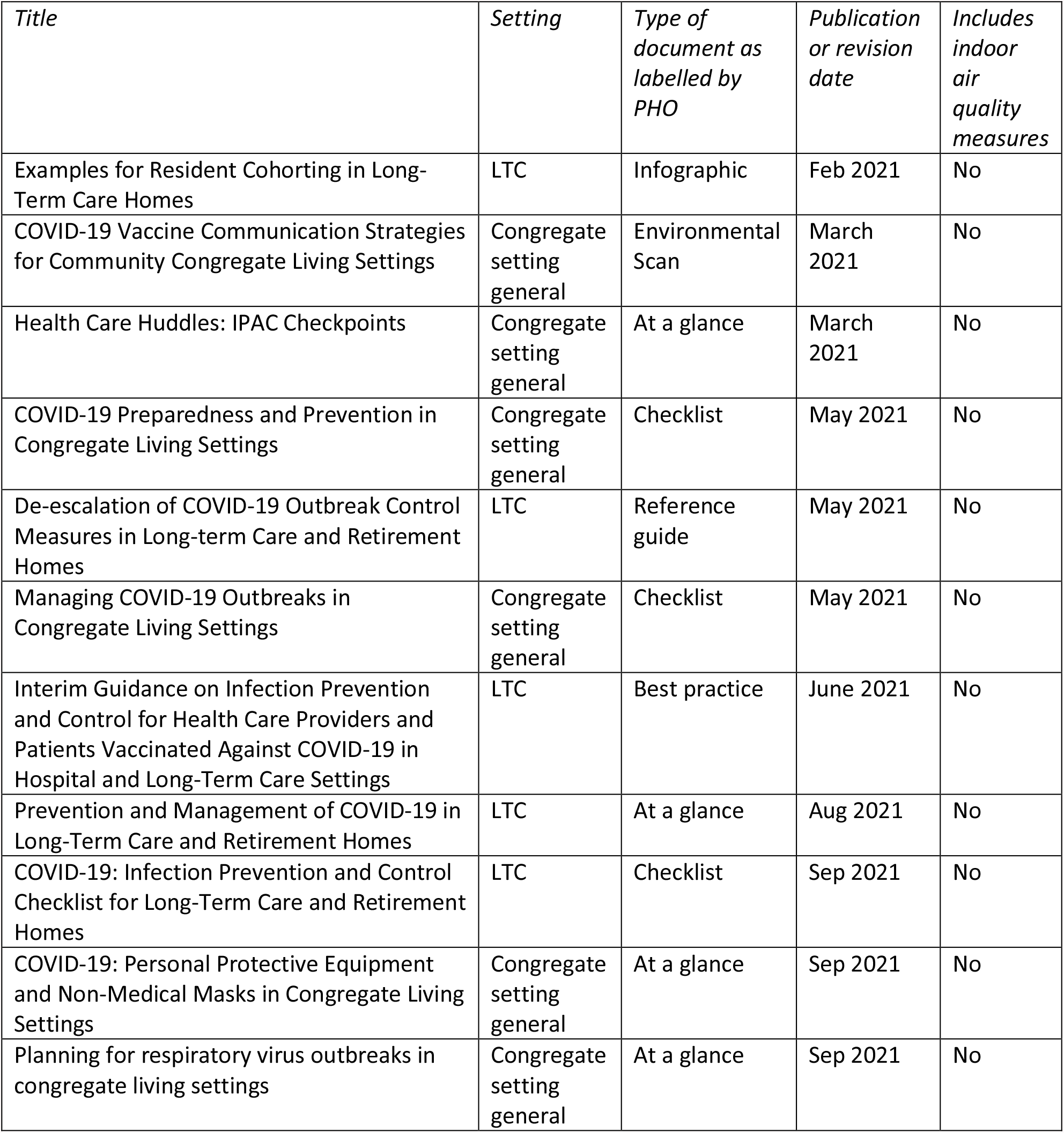
General and topic-specific COVID-19 guidance developed specifically for long-term care (LTC) and congregate settings, published or revised between January and October, 2021 (n = 11).

### Comparing COVID-19 checklists

The nine checklists included in this category have varying foci. Some are focused on planning and preparation, others on COVID-19 outbreaks and still others on daily operations (see Table 2). All, however, include sections on general infection prevention and control measures, and are interactive documents designed to direct the activities of the reader. We focused on the most recent revisions of each checklist published up to October 29, 2021.

**Table 2.** Setting-specific COVID-19 checklists from PHO (n = 9), including two checklists for congregate settings and one for long-term care (LTC) homes, published or revised between January and October, 2021.

| *Title* | *Setting* | *Publication or revision date* | *Includes indoor air quality measures* |
| --- | --- | --- | --- |
| COVID-19 Preparedness and Prevention in Congregate Living Settings | Congregate setting general | May 2021 | No |
| Daily Camp Operations: COVID-19 Preparedness and Prevention for Day Camps | Day camps | May 2021 | Yes |
| Managing COVID-19 Outbreaks in Congregate Living Settings | Congregate setting general | May 2021 | No |
| Pre-camp Planning: COVID-19 Preparedness and Prevention for Day Camps | Day camps | May 2021 | Yes |
| Daily Camp Operations: COVID-19 Preparedness and Prevention for Overnight Camps | Overnight camp | June 2021 | Yes |
| Pre-camp Planning: COVID-19 Preparedness and Prevention for Overnight Camps | Overnight camp | June 2021 | Yes |
| COVID-19 Preparedness and Prevention in Elementary and Secondary (K-12) Schools | Schools | Sep 2021 | Yes |
| Infection Prevention and Control Key Principles for Clinical Office Practice During the COVID-19 Pandemic | Clinical offices | Sep 2021 | Yes |
| COVID-19: Infection Prevention and Control Checklist for Long-Term Care and Retirement Homes | LTC | Sep 2021 | No |

We identified two COVID-19 checklists for **congregate settings** (both revised May, 2021) and one for **long-term care and retirement homes** (revised, September, 2021). These resources do not contain references to IAQ measures. (Please note, these checklists are also included in our primary sample as analyzed in the previous section.)

We identified one COVID-19 checklist for **schools** (revised, September, 2021). This checklist contains a dedicated section on IAQ, which includes references to HVAC systems, natural ventilation and portable air filters.

We identified one COVID-19 checklist for **clinical office practice** (revised September, 2021). This checklist contains a dedicated section on HVAC systems listing specific HVAC standards. It suggests that if HVAC systems do not meet these standards, facility managers should consider mitigation measures such as open windows or air filtration.

We identified two COVI9-19 checklists for **day camps** (both published May, 2021), and two for **overnight camps** (both published June, 2021). Checklists focused on pre-camp planning suggest the need to designate a well-ventilated area for isolation. Checklists focused on daily operations suggest opening windows and tent flaps to increase natural ventilation, and ensuring HVAC systems (if applicable) are regularly maintained.

Please see Table 2 for all checklists.

### Additional guidance

During our initial search, we identified slide presentations for two 2021 webinars listed on the relevant pages of PHO’s website. As we chose to focus on written guidance, we did not include these webinars in our sample. We did, however, attempt to watch both. The first was not posted on the PHO website, and we could not find it through a Google search. The second was not posted on the PHO website, but we did find it through a Google search, posted on YouTube. Although the second webinar does have a slide mentioning HVAC, the discussion is focused on how to plan for infection prevention and control issues during renovations such as exposing residents to hazards, or interrupting essential facility functions.

In addition, during our initial search, we identified written documents (n. 6) posted to the “congregate settings” and “long-term care” pages of PHO’s website that were not designed specifically for long-term care or congregate settings, but rather for general or health care settings. Please see Table 3 for a list of these documents. Three of these documents discuss IAQ measures. One is a general guidance document on HVAC in buildings during COVID-19, one is a “Q and A” about ultra-violet disinfection, and one is interim guidance for health care settings that includes a recommendation about reviewing HVAC systems (this document includes long-term care homes in its definition of health care settings).

**Table 3.** Additional documents posted to the “congregate settings” and “long-term care” pages but not designed specially for long-term care or congregate settings (n = 6).

| *Title* | *Setting* | *Type* | *Publication or revision date* | *Includes indoor air quality measures* |
| --- | --- | --- | --- | --- |
| Heating, Ventilation and Air Conditioning (HVAC) Systems in Buildings and COVID-19 | Public | Focus on | March 2021 | Yes |
| Coronavirus Disease 2019 (COVID-19) Cleaning and Disinfection for Public Settings | Public | Fact sheet | July 2021 | No |
| Key Elements of Environmental Cleaning in Healthcare Settings | Health care | Fact sheet | July 2021 | No |
| Ultraviolet (UV) light disinfection for SARS-CoV-2 | Public | FAQ | July 2021 | Yes |
| Interim Guidance for Infection Prevention and Control of SARS-CoV-2 Variants of Concern for Health Care Settings | Health care | Best practice | Aug 2021 | Yes |
| Key features of influenza, SARS-CoV-2 and Other Common Respiratory Viruses | Public | At a glance | Sep 2021 | No |

## Discussion

PHO has omitted references to IAQ measures from its public, written guidance specifically designed for long-term care and congregate settings. This omission, however, does not appear across all of PHO’s materials. For example, PHO includes limited references to IAQ measures in its COVID-19 checklists for schools, summer camps and clinical offices. In addition, in March 2021, PHO published detailed guidance related to HVAC systems and COVID-19 for general audiences,^43^ followed by an “FAQ” on ultra-violet disinfection in July, 2021.^44^ PHO has also hosted webinars specifically focused on indoor air quality.^45,46^ Finally, in May, 2021, PHO published an evidence synthesis focused on droplet and aerosol transmission. This evidence synthesis concludes with recommendations for reducing transmission, one of which is as follows:

> Ensuring that ventilation systems are well-maintained and optimized with the support of professionals according to relevant recommendations (e.g., from American Society of Heating, Refrigerating and Air-Conditioning Engineers) and/or using outdoor environments whenever possible.^47p.15^

We will not speculate as to why PHO recommends indoor air quality measures in COVID-19 checklists for some settings and not others, or why its own detailed recommendations around HVAC systems are not referenced in its guidance for long-term care or congregate settings. Rather, we suggest that future research focus on accountability, with an emphasis on regulatory and legal remedies.

The issues outlined in this paper were explored 15 years ago by the SARS Commission report, through which runs a tone of desperation and anger in response to the preventable loss of human life. Report author Archie Campbell pleads with decision-makers in Ontario not to wait for certainty to protect people’s lives during the next pandemic, writing, “If the Commission has one single take-home message it is the precautionary principle that safety comes first, that reasonable efforts to reduce risk need not await scientific proof.”^1p.13^

In the case of airborne transmission of COVID-19, however, we do have scientific proof, as outlined in this paper. There are no pretexts left upon which to omit measures such as ventilation, filtration and ultra-violet disinfection from COVID-19 infection prevention and control. It is imperative that all Ontario facilities receive updated public health guidance in the short term. This is particularly urgent in the context of facilities where structures such as colonization, white supremacy and ableism determine who is compelled to live there, and who is not.

## Limitations

It’s possible that some relevant resources on PHO’s website are not listed on their “congregate living” and “long-term care” pages. As a result, it is possible that we have not captured all PHO’s public, written guidance for congregate settings. Searches such as “COVID-19” and “long-term care” or “COVID-19” and “congregate settings” on PHO’s website generate several dozen entries, often for resources that overlap in content, and with little indication as to which type of document may be the most comprehensive or definitive. In addition, Google searches may turn up additional resources.

We assumed, however, that workers, facility managers and inspectors would not sift through search results on PHO’s website, or conduct Google searches, but rather focus on guidance, and especially the checklists, posted on relevant areas of the organization’s website.

We also assumed guidance produced by PHO for long-term care and congregate settings has some impact on long-term care and congregate settings. This study, however, did not explore the extent to which PHO guidance influences those responsible for health and safety in specific facilities. An earlier pilot review did find, however, that PHO’s guidance for congregate settings heavily influenced advice provided by some local Public Health Units to facilities such as shelters.^41^ Future research may wish to explore where facility operators go for guidance they consider to be useful, definitive and/or binding.

In addition, we did not look at all recent guidance for our comparator settings of schools, summer camps and clinical offices. It is possible that the guidance we looked at is not representative of what PHO is generally sharing with these settings. We made the assumption, however, that COVID-19 checklists were likely to be representative documents, given that they are designed to coordinate action by those responsible for health and safety.

We should also note that while COVID-19 checklists for schools, summer camps and clinical offices include some IAQ measures, this guidance could also be classified as “sub-standard.” For example, none of these checklists recommend portable air filtration, bathroom fans, protocols for flushing room air between cohorts, or UV disinfection. As a result, comparisons generated by our study may serve to overstate the quality of the guidance provided to settings such as schools.

Finally, we do not have comprehensive insight into the current state of IAQ practices in long-term care and congregate settings in Ontario. We do, however, have partial insight, as some on our team are regularly consulted by managers and workers from congregate settings, who, in the absence of formal IAQ guidance from public health authorities, seek alternative expertise.

## Conclusion

PHO’s written, public COVID-19 guidance for long-term care and congregate settings has omitted basic IAQ measures such as portable air filtration, HVAC optimization, bathroom fans that exhaust to the outside, clearing the air between cohorts, and UV disinfection. These measures, when implemented and maintained with expert advice, generate virtually no health risks, and are very likely to reduce transmission of COVID-19 and other respiratory illnesses. By omitting IAQ measures from its COVID-19 guidance, PHO has generated no health benefit to residents or workers, but rather increased their risk of illness and death.

PHO’s failure to provide high-quality guidance to long-term care homes and congregate settings has particular implications for health equity. Inter-related structures such as colonization, white supremacy and ableism determine the populations that are compelled by the Canadian state and its institutions to live in settings such as shelters, detention centers, group homes and long-term care homes. These same structures determine which populations are generally able to avoid these facilities. As a result, omitting IAQ measures from COVID-19 guidance to long-term care and congregate settings exacerbates health inequities in Ontario.

PHO is mandated by legislation to share scientific and technical advice during infectious disease outbreaks, and to reduce health inequities. In the context of its written, public, COVID-19 guidance to long-term care homes and congregate settings, PHO has not met this mandate. As a result, we recommend that future research focus on the question of accountability, with an emphasis on regulatory and legal remedies.

## Data Availability

Most data associated with this study are contained in the manuscript and tables. All data associated with this study are available upon reasonable request to the authors.

## Acknowledgements

Thank you to Jessica Demeria, Melissa Goldstein and Kate Francombe-Pridham for generously sharing your knowledge and insight during the early stages of this work. Thank you to Karissa Avignon for your thoughtful and painstaking review of our data and conclusions.

## Footnotes

1 In December, 2021, the Ontario Ministry and Long-Term Care updated its COVID-19 directive to include the routine use of N95 masks when treating patients who have or who are suspected to have COVID-19. This change is reflected in a guidance document published by PHO on December 23, 2021 for long-term care and retirement homes, “COVID-19: Self-Assessment Audit Tool for Long-term Care Homes and Retirement Homes.” This document also includes one reference to an IAQ measure, “HVAC systems are functioning properly (check with facility manager).”

